# Characteristics associated with household transmission of SARS-CoV-2 in Ontario, Canada

**DOI:** 10.1101/2020.10.22.20217802

**Authors:** Lauren A. Paul, Nick Daneman, Kevin A. Brown, James Johnson, Trevor van Ingen, Eugene Joh, Sarah E. Wilson, Sarah A. Buchan

## Abstract

**BACKGROUND:** Within-household transmission of SARS-CoV-2 infection has been identified as one of the main sources of spread of COVID-19 after lockdown restrictions and self-isolation guidelines are implemented. Secondary attack rates among household contacts are estimated to be five to ten times higher than among non-household contacts, but it is unclear which individuals are more prone to transmit infection within their households.

**METHODS:** Using address matching, a cohort was assembled of all laboratory-confirmed cases of COVID-19 residing in private households in Ontario, Canada. Descriptive analyses were performed to compare characteristics of cases in households that experienced secondary transmission versus those that did not. Logistic regression models were fit to determine index case characteristics and neighbourhood characteristics associated with transmission.

**FINDINGS:** Between January and July, 2020, there were 26,152 cases of COVID-19 residing in 21,226 households. Longer testing delays (≥5 days versus 0 days OR=3·02, 95% CI: 2·53 - 3·60) and male sex (OR=1·28, 95% CI: 1·18 - 1·38) were associated with greater odds of household secondary transmission, while being a healthcare worker (OR=0·56, 95% CI: 0·50 - 0·62) was associated with lower odds of transmission. Neighbourhoods with larger average economic family size and a higher proportion of households with multiple persons per room were also associated with greater odds of transmission.

**INTERPRETATION:** It is important for individuals to get tested for SARS-CoV-2 infection as soon as symptoms appear, and to isolate away from household contacts; this is particularly important in neighbourhoods with large family sizes and/or crowded households.

**FUNDING:** This study was supported by Public Health Ontario.

**Research in Context:** *EVIDENCE BEFORE THIS STUDY:* We searched PubMed and Google Scholar up to September 3, 2020 to identify individual-level cohort studies or meta-analyses on household transmission of COVID-19. We used the search terms (“COVID” OR “SARS-CoV-2”) AND (“household” [Title]), and also reviewed the reference lists of any studies found during the search to identify additional studies. We considered studies that reported secondary attack rates and/or other measures of association (i.e., relative risk, odds ratio, or hazard ratio) for household transmission. We did not consider any modelling studies, studies that focused specifically on children, or small case studies that included less than three households. The search returned 53 studies, of which 51 were included in three meta-analyses. Pooled household secondary attack rates from the three meta-analyses were 19%, 27%, and 30%; secondary attack rates in households were estimated to be five to ten times as high as in non-household settings. Most studies were conducted in Asia and identified households from contact tracing, with individual studies reporting on fewer than 6000 households. Most studies did not consider households with no secondary transmission, and focused on a limited set of secondary case characteristics.

*ADDED VALUE OF THIS STUDY:* We applied an address matching algorithm, which identified 21,226 private households of laboratory-confirmed cases of COVID-19 in Ontario, Canada. Ontario has the advantage of a universal healthcare system and population-wide data for the entire province. To our knowledge, this study contains the largest number of private households with at least one confirmed case of COVID-19. We compared a variety of individual- and neighbourhood-level characteristics of households with and without secondary transmission. We also applied logistic regression models to determine index case characteristics associated with transmission, which gave important insights into factors that may help reduce secondary transmission in households.

*IMPLICATIONS OF ALL THE AVAILABLE EVIDENCE:* Findings from this study and existing evidence suggest that testing delays and household crowding play important roles in whether household secondary transmission occurs. Odds of household transmission may be reduced by cases seeking testing as soon as symptoms appear, and self-isolating outside the home or in a room alone if possible. These strategies may be considered by public health officials to reduce household transmission and mitigate local spread of COVID-19. Future research should further investigate the role of children and youth in household transmission.

## Introduction

Transmission and acquisition of SARS-CoV-2 infection has become an active area of COVID-19 research since person-to-person transmission was confirmed at the beginning of 2020.^1,2^ In many countries, the primary source of acquisition of SARS-CoV-2 infection has transitioned from travel-related transmission early in the pandemic, to local transmission as countries implemented travel restrictions to reduce imported infections. Within-household transmission in particular has been highlighted as an important source of COVID-19 transmission for communities.^3–7^ The shift to household transmission has arisen due to the fact that many public health measures, ranging from teleworking to full lockdowns, encourage individuals to spend more time at home where there is increased duration and intensity of contact among household members.^4,5^ However, it is unclear which individuals are more likely to transmit infection within their households.

Existing individual-level observational studies of household transmission typically included household contacts identified through contact tracing.^4–6,8–10^ These studies have estimated secondary attack rates among household contacts to be five to ten times as high as in non-household settings.^4,6^ Most of these studies were conducted in Asia, included smaller numbers of households, and/or did not compare to households where no secondary transmission occurred. Many also focused on the characteristics of the acquirers of infection (secondary cases) rather than the characteristics of the transmitters of infection (index cases) in the household.

Using address matching, we sought to identify all households with confirmed SARS-CoV-2 infections from Ontario, Canada between January and July, 2020. We were interested in comparing characteristics of cases in households that experienced secondary transmission (i.e., additional laboratory-confirmed cases following the index case) versus those that did not, and also sought to determine individual- and neighbourhood-level characteristics of index cases associated with transmission. This work may help inform future public health strategies to reduce within-household transmission during the ongoing pandemic.

## Methods

### Study population

We assembled a cohort of all confirmed cases of COVID-19^11^ reported in Ontario, Canada’s most populous province (14 million residents), among residents of private households from January 1, 2020 to July 28, 2020. We identified confirmed cases of COVID-19 using data from provincial reportable disease systems entered by local public health units.^12^ We obtained ethics approval from Public Health Ontario’s Research Ethics Board.

### Identification of private households

Private households were defined as any residences not identified as congregate in nature, such as homeless shelters or long-term care homes. Individual houses and apartments/suites within multi-unit dwellings (e.g., apartment buildings) were considered private households. For address matching, we applied a natural language processing algorithm using Python’s sklearn library to identify unique households that contained at least one COVID-19 case.^13^ Briefly, we broke down cases’ whole address fields (including street address, city, and postal code) and found a closest match in a master list of addresses, containing congregate facilities and previously identified households. This match was then validated by checking for exact matches in the numerical portion of the address field. For unmatched addresses, we again used a natural language processing algorithm to identify duplicates, and added unique addresses to the master list for future comparisons. We excluded any cases whose address matched a known congregate facility or who had a risk factor flag for residing in a congregate setting in provincial reportable disease systems. We also examined addresses that were matched with apartment buildings in the master list for suite information, and excluded cases missing suite information as we were unable to determine conclusively whether these individuals resided in the same suite as others in the building. We excluded any cases with missing or incomplete address information.

### Outcomes

The primary outcome of interest was any secondary transmission within a household, defined as cases that occurred 1-14 days after the index case of the household.^8,10,14^ We used each case’s symptom onset date as the date of comparison, or their specimen collection date if symptom onset date was unavailable, and excluded the rare cases (0·5%) that lacked information on both dates. We excluded households with multiple cases on the index date (“index clusters”) from the cohort as they would present challenges for estimating the predictive value of individual-level characteristics. We also considered two secondary outcomes of interest: household transmission to older adults (≥60 years), and household transmission to severe cases (ICU admission or death).

### Individual-level characteristics

We considered a variety of individual-level and neighbourhood-level covariates in our analyses that were hypothesized to influence household transmission. At the individual level, we obtained information on each case’s age, sex, and health region of residence (Supplementary Table S1). Furthermore, we included covariates for case month (January–July), employment as a healthcare worker, high risk status (≥60 years of age, immunocompromised, had cardiovascular-related health issues, or had chronic obstructive pulmonary disease (COPD)), and association with a known COVID-19 outbreak outside the home (e.g., association with a workplace outbreak or long-term care home outbreak).^11^

We also considered three delay metrics for each case: (1) the delay between the case’s symptom onset and when their specimen was collected by a healthcare provider (testing delay); (2) the delay between specimen collection and report of a positive test result to the local health unit (reporting delay); and (3) the delay between test report and when the health unit begins entering the case into a reportable disease system for provincial notification (data entry delay). For the testing delay metric, we additionally separated out cases who were missing symptom onset date (thus specimen collection date was used) and did not have any COVID-19 symptoms flagged in provincial disease reporting systems. We excluded cases that were missing symptom onset date but had COVID-19 symptoms flagged from all analyses.

We did not have any information on the total number of residents of each household. However, we were able to adjust for several characteristics related to the average size and composition of households at the neighbourhood level.

### Neighbourhood-level characteristics

At the neighbourhood level, we had information available from 2016 Canadian census records (98·4% response rate^15^). The Canadian census is a mandatory questionnaire that collects extensive information from each of the 15·4 million dwellings across Canada, with all dwellings reporting household composition, and a 25% sample completing a more detailed long-form questionnaire.^15^ We linked neighbourhood characteristics at the aggregate dissemination area level, which divides the country into areas with populations between 5,000 and 15,000 persons, on average. These included characteristics such as the average economic family size, proportion of households with multiple persons per room, proportion of multi-family households, and urban/rural status (see Supplementary Definitions). A full list of the neighbourhood characteristics is found in Table 3.

### Statistical analysis

We applied logistic regression models to obtain both unadjusted and adjusted odds ratios (OR) and 95% confidence intervals (CI) for the associations between covariates and the odds of secondary transmission within a household. We also carried out several descriptive analyses to compare the characteristics of index cases, secondary cases, and cases that were not involved in any household transmission. We explored the breakdown of these groups by outcome severity (i.e., hospital admission, ICU admission, death, or no serious or severe outcome) and examined the direction of transmission by age group and high risk status. We assessed the distribution of the number of days between symptom onset dates for index cases and secondary cases (serial interval).

In sensitivity analyses we adjusted the definition of household transmission to be (1) cases that occurred 2-14 days after the index case (more specific) or (2) cases that occurred 1-28 days after the index case (more sensitive). We also restricted the analysis to households with an index case date on or after May 29; testing approaches expanded as of May 29, which may have improved the ability to identify secondary transmission in households.^16^

### Role of the funding source

This study was supported by Public Health Ontario. The authors had full access to all data in the study and accept responsibility for the decision to submit for publication.

## Results

As of July 28, 2020, there were 38,984 confirmed cases of COVID-19 reported in Ontario. After removing cases based on our inclusion criteria, we were left with 26,152 cases residing in private households, of which 18,159 cases were from households with no secondary transmission and 7,993 cases were from households with secondary transmission (Figure 1). Of the 3,067 index cases from households with secondary transmission, the median number of secondary cases in the same household was one (25^th^ percentile=one case, 75^th^=two cases, 90^th^=three cases). The average age of the cohort was approximately 44 years and 53% were female.

**Figure 1.**
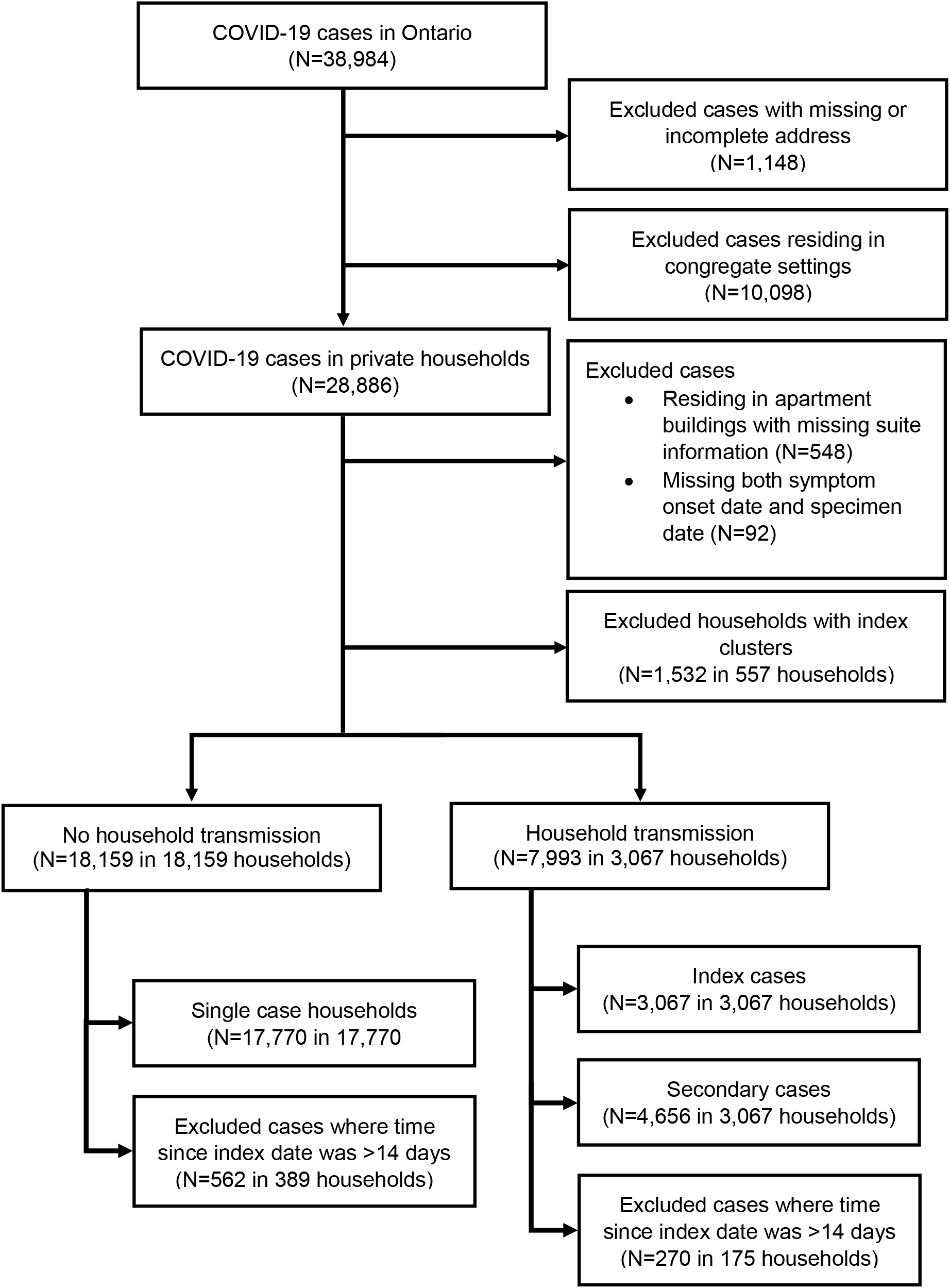
Flow diagram of study cohort.

### Timing and direction of transmission

The median serial interval from index case to secondary case was four days (25^th^ percentile=two days, 75^th^=seven days, 90^th^=ten days) (Supplementary Figure S1). For the direction of transmission from index cases to secondary cases, individuals in the 20-59 year age group and low risk individuals were both the most frequent transmitters and most frequent acquirers of SARS-CoV-2 infections within households (Figure 2). Transmissions to secondary cases in different age or risk groups than the index case were less frequent.

**Figure 2.**
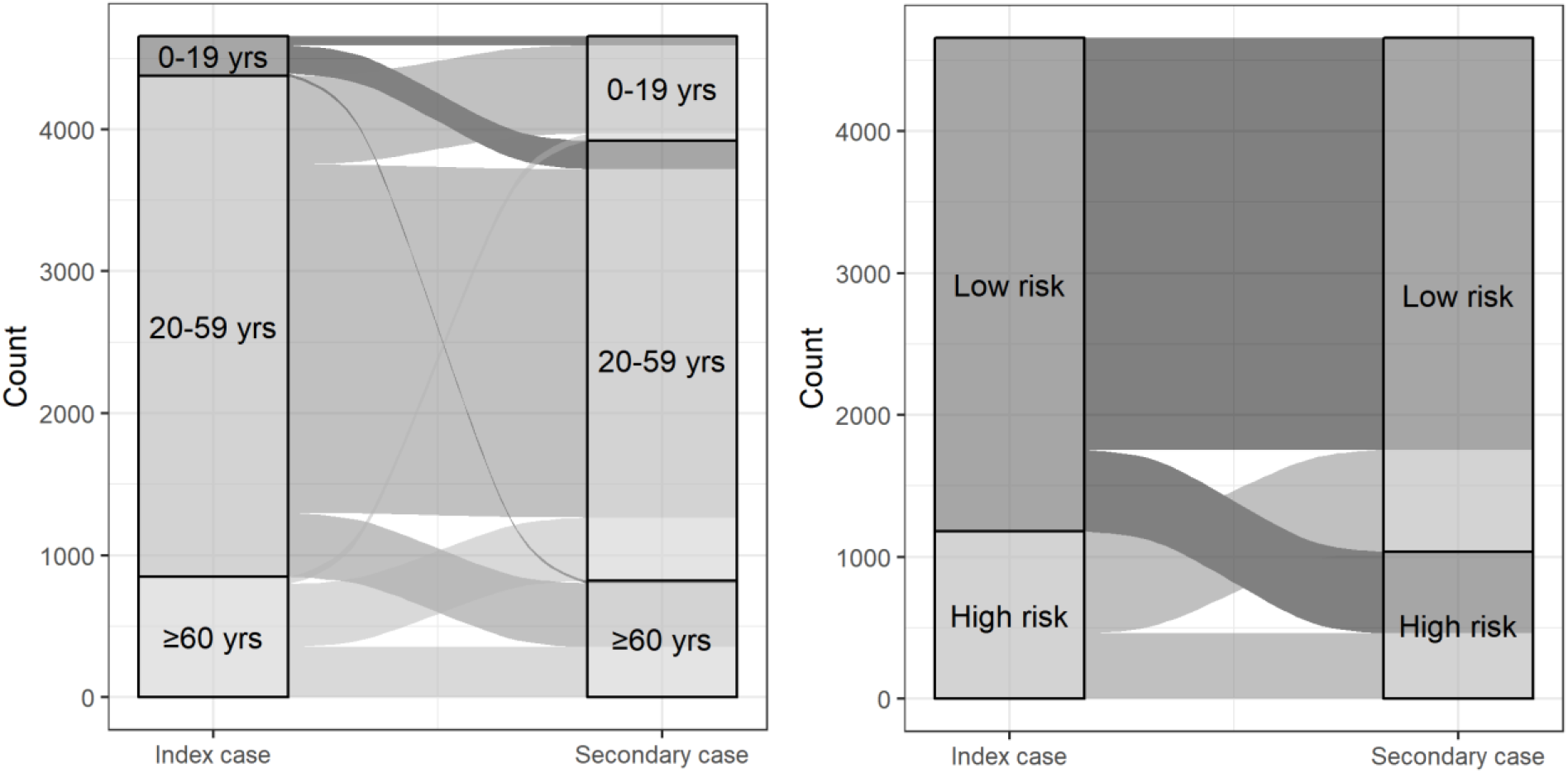
Direction of transmission from index case to secondary case by (A) age group and (B) high risk status. The line represents the direction of transmission from index case to secondary case. The shade of the line represents the age group or risk group of the index case. The width of the line is proportional to the frequency of transmission between index and secondary cases in their respective age or risk groups. Yrs=years.

### Comparison of individual-level characteristics

Compared to index cases with no household transmission, index cases with household transmission were less likely to be healthcare workers or associated with a known COVID-19 outbreak (Table 1). However, they were more likely to be male and had median testing delays that were twice as long as index cases without household transmission (four days versus two days). There was no difference in median reporting delay or data entry delay for the two groups.

**Table 1.** Characteristics of index cases in households with no household transmission compared to index cases of households with household transmission

|  | Index cases with<br>no household transmission<br>(N=18,159) | Index cases with<br>household transmission<br>(N=3,067) |
| --- | --- | --- |
| Sex (N, %) |  |  |
| Female | 9,890 (54.5) | 1,464 (47.7) |
| Male | 8,214 (45.2) | 1,595 (52.0) |
| Age, years (median, IQR) | 45 [31, 58] | 46 [31, 57] |
| Age group (N, %) |  |  |
| <10 years | 164 (0.9) | 26 (0.8) |
| 10-19 years | 536 (3.0) | 127 (4.1) |
| 20-29 years | 3,387 (18.7) | 523 (17.1) |
| 30-39 years | 3,169 (17.5) | 481 (15.7) |
| 40-49 years | 3,256 (17.9) | 571 (18.6) |
| 50-59 years | 3,711 (20.4) | 726 (23.7) |
| 60-69 years | 2,271 (12.5) | 404 (13.2) |
| 70-79 years | 972 (5.4) | 138 (4.5) |
| ≥80 years | 692 (3.8) | 70 (2.3) |
| High risk (≥60 years, immunocompromised,<br>cardiovascular, COPD) (N, %) | 5,066 (27.9) | 844 (27.5) |
| Outbreak-associated* (N, %) | 4,901 (27.0) | 540 (17.6) |
| Healthcare worker (N, %) | 4,916 (27.1) | 517 (16.9) |
| Month reported (N, %) |  |  |
| January | 1 (0.0) | 1 (0.0) |
| February | 8 (0.0) | 3 (0.1) |
| March | 2,009 (11·1) | 312 (10·2) |
| April | 6,374 (35·1) | 945 (30·8) |
| May | 4,978 (27·4) | 989 (32·2) |
| June | 2,931 (16·1) | 528 (17·2) |
| July | 1,858 (10·2) | 289 (9·4) |
| Region (N, %) |  |  |
| Toronto | 6,292 (34·6) | 1,025 (33·4) |
| Central East | 5,891 (32·4) | 1,236 (40·3) |
| Central West | 2,445 (13·5) | 343 (11·2) |
| Eastern | 1,569 (8·6) | 231 (7·5) |
| Northern | 199 (1·1) | 32 (1·0) |
| South West | 1,763 (9·7) | 200 (6·5) |
| Testing delay†, days (median, IQR) | 2 [0, 6] | 4 [2, 8] |
| Testing delay distribution† (N, %) |  |  |
| No symptoms‡ | 2,883 (16·2) | 131 (4·3) |
| <0 days§ | 963 (5·4) | 60 (2·0) |
| 0 days | 1,745 (9·8) | 164 (5·4) |
| 1 day | 1,955 (11·0) | 349 (11·5) |
| 2 days | 1,958 (11·0) | 341 (11·2) |
| 3 days | 1,560 (8·8) | 327 (10·8) |
| 4 days | 1,230 (6·9) | 276 (9·1) |
| ≥5 days | 5,529 (31·0) | 1,390 (45·8) |
| Reporting delay, days (median, IQR) | 2 [1, 3] | 2 [1, 3] |
| Reporting delay distribution (N, %) |  |  |
| <0 days | 312 (1·7) | 43 (1·4) |
| 0 days | 1,165 (6·4) | 200 (6·5) |
| 1 day | 5,931 (32·8) | 1,038 (34·0) |
| 2 days | 5,276 (29·2) | 926 (30·3) |
| 3 days | 2,414 (13·4) | 390 (12·8) |
| 4 days | 1,010 (5·6) | 188 (6·2) |
| ≥5 days | 1,966 (10·9) | 271 (8·9) |
| Data entry delay, days (median, IQR) | 0 [0, 1] | 0 [0, 1] |
| Data entry delay distribution (N, %) |  |  |
| <0 days | 1,065 (5·9) | 173 (5·6) |
| 0 days | 11,050 (60·9) | 1,852 (60·4) |
| 1 day | 3,699 (20·4) | 696 (22·7) |
| 2 days | 824 (4·5) | 132 (4·3) |
| 3 days | 457 (2·5) | 80 (2·6) |
| 4 days | 286 (1·6) | 32 (1·0) |
| ≥5 days | 778 (4·3) | 102 (3·3) |

\*Cases associated with a public health declared outbreak outside the home.
†269 cases were excluded that had COVID-19 symptoms flagged in provincial reportable disease systems but were missing symptom onset date.
‡Cases with no symptoms were defined as cases that were missing symptom onset date (thus specimen collection date was used) and did not have any COVID-19 symptoms flagged in provincial reportable disease systems.
§Cases with a testing delay of <0 days were those who were tested prior to the onset of their symptoms.

We also compared the characteristics of index cases and secondary cases, and found that secondary cases had shorter median testing delays than index cases (Supplementary Table S2). They were also less likely to have serious or severe outcomes (Supplementary Table S3).

### Associations with delay metrics

From adjusted logistic models, we observed increased odds of any household transmission with longer testing delays for the index case compared to 0-day (i.e., the individual was tested on the same day as their symptom onset) testing delays (ORs: 1-day delay=2·02, 2-day delay=1·96, 3-day delay=2·36, 4-day delay=2·64, ≥5-day delay=3·02) (Figure 3, Supplementary Table S4). Individuals with no symptoms flagged in provincial reportable diseases systems had lower odds of any household transmission (0·48, 95% CI: 0·38 - 0·61). This trend was similar in our models for household transmission to older adults and to cases with severe outcomes. Conversely, there were no notable trends for increased odds of household transmission with reporting delays or data entry delays.

**Figure 3.**
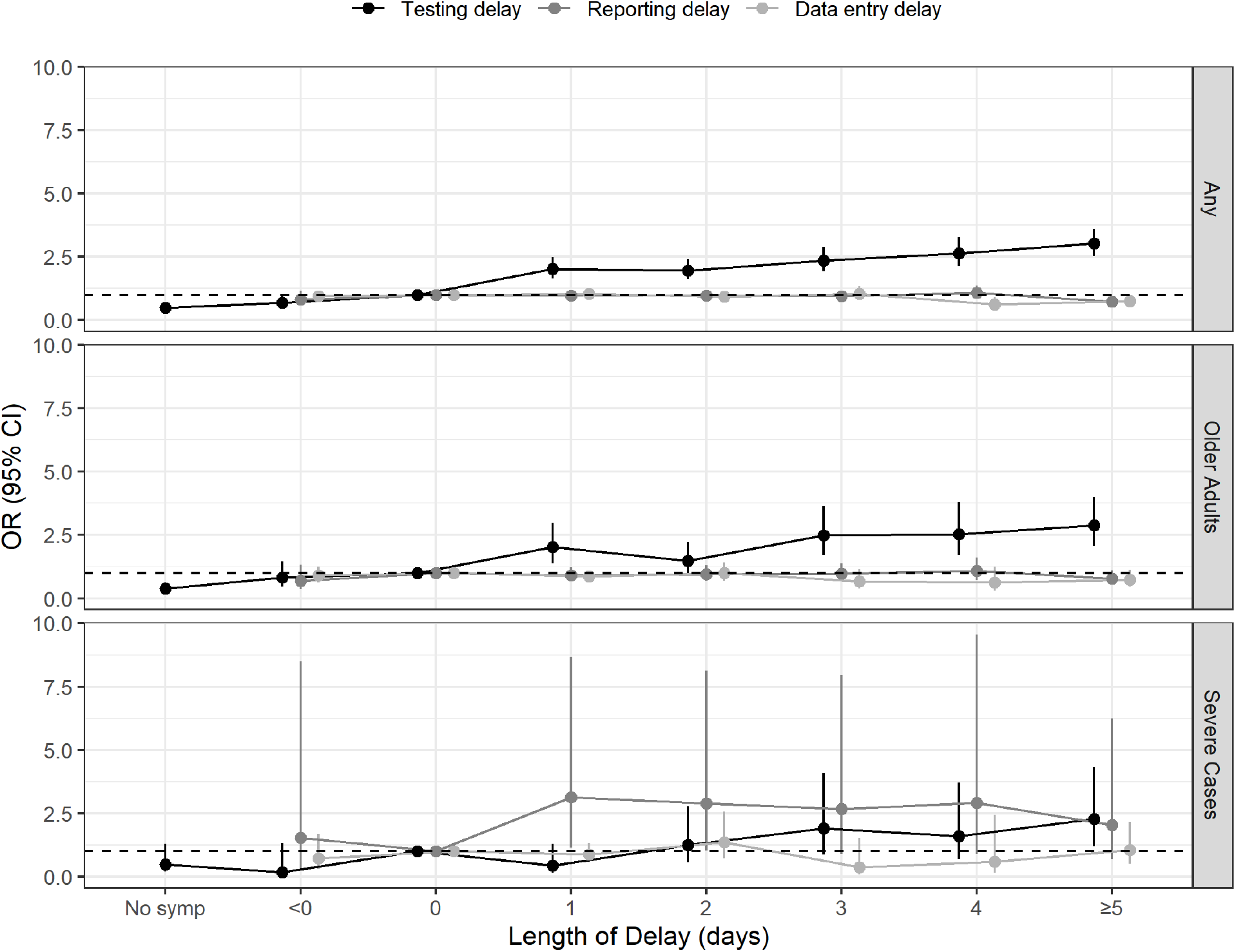
Adjusted odds ratios and 95% confidence intervals for the associations between index case delay metrics and odds of household transmission 269 cases were excluded from the testing delay models that had COVID-19 symptoms flagged in provincial reportable disease systems but were missing symptom onset date. Cases with no symptoms were defined as cases that were missing symptom onset date (thus specimen collection date was used) and did not have any COVID-19 symptoms flagged in provincial reportable disease systems. Cases with a testing delay of <0 days were those who were tested prior to the onset of their symptoms. Horizontal line at OR=1 indicating no association. No symp=no symptoms.

### Associations with other individual-level characteristics

Male index cases had higher odds of any household transmission (1·28, 95% CI: 1·18 - 1·38) or transmission to older adults (1·19, 95% CI: 1·02 - 1·38) compared to female index cases, and older (≥60 years) and younger (20-49 years) index cases had lower odds of any household transmission compared to the 50-59 year reference group (Table 2). We observed increased odds of household transmission if the index case was high risk (1·14, 95% CI: 0·97 - 1·34), and decreased odds if the index case was a healthcare worker (0·56, 95% CI: 0·50 - 0·62) or was associated with a known outbreak (0·61, 95% CI: 0·55 - 0·68). There were also some trends for decreased odds of transmission from May to July.

**Table 2.**
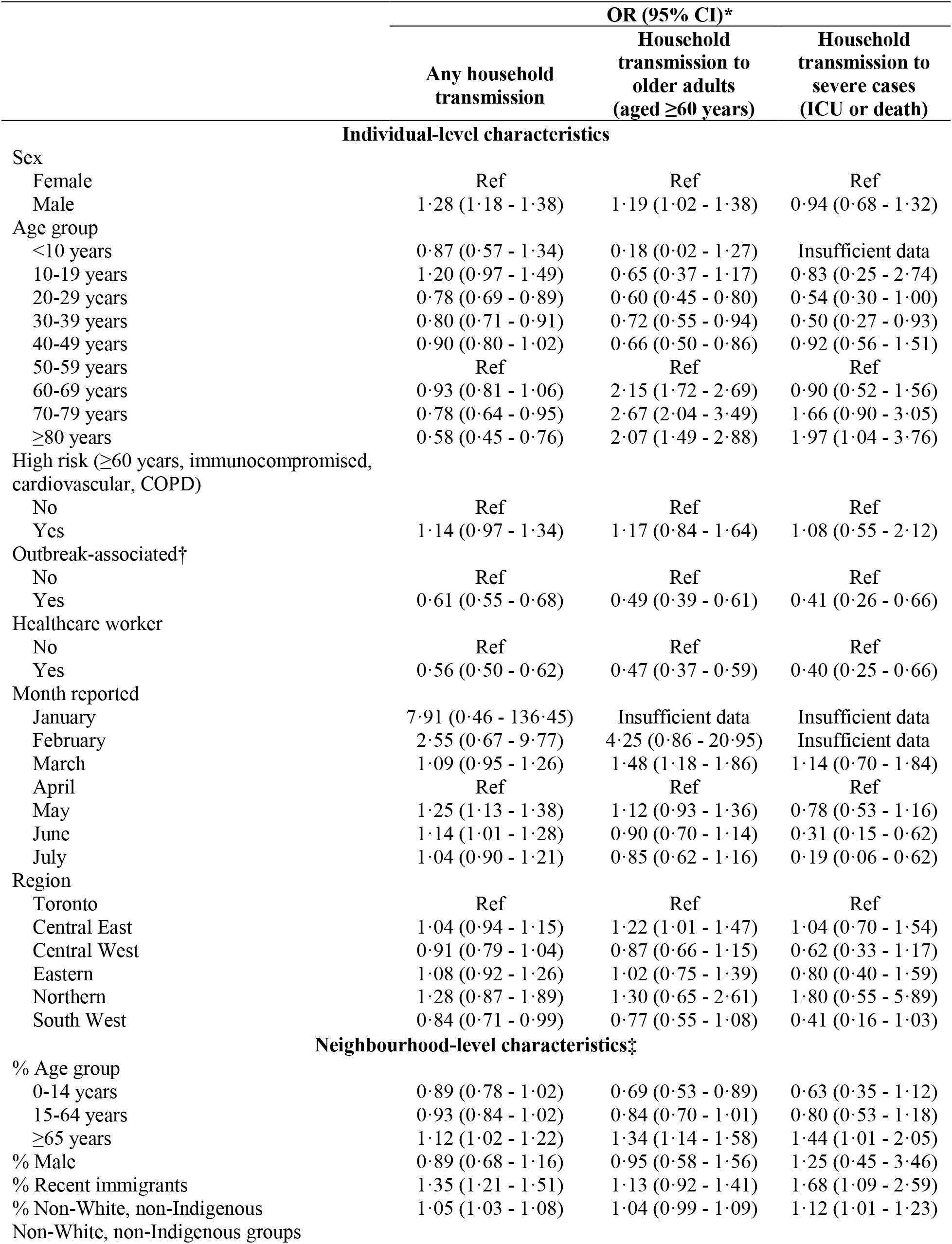
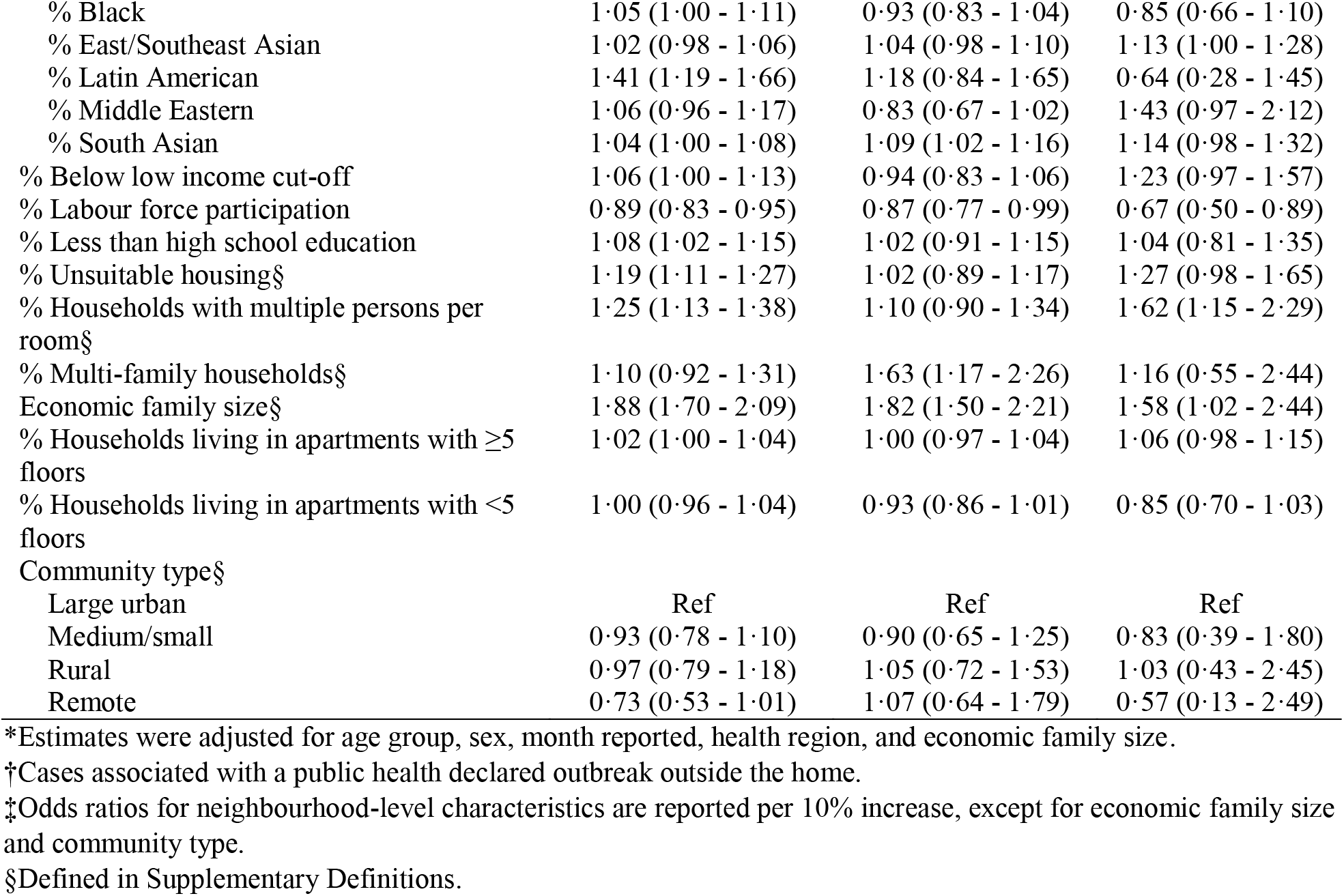
Adjusted odds ratios and 95% confidence intervals for the associations between index case characteristics and odds of household transmission

### Associations with neighbourhood-level characteristics

The strongest associations observed for household transmission were in neighbourhoods with larger average economic family size (1·88 per person increase, 95% CI: 1·70 - 2·09 per person increase) or neighbourhoods with a higher proportion of households with multiple persons per room (1·25 per 10% increase, 95% CI: 1·13 - 1·38). We also observed increased odds for neighbourhoods with a higher proportion of multi-family households; this was a particularly strong predictor of transmission to older adults (1·63 per 10% increase, 95% CI: 1·17 - 2·26). Additionally, odds of transmission were higher for neighbourhoods with a higher proportion of individuals in the ≥65 year age group; individuals below the low income cut off; individuals with less than high school education; unsuitable housing; recent immigrants; non-White, non-Indigenous groups; and apartments with five or more floors (Table 2). Odds were lower for neighbourhoods with a higher proportion of individuals participating in the labour force, as well as more remote areas compared to large urban areas.

### Sensitivity analysis

We compared the estimates from our primary outcome model with those produced in our three sensitivity analyses (i.e., household transmission 2-14 days after index cases and 1-28 days after index cases, and index case dates on or after May 29), and found that our associations were robust (Supplementary Table S5). Notably, longer testing delays continued to display strong trends towards increased odds of household transmission. Larger average economic family size and a higher proportion of households with multiple persons per room also continued to exhibit the strongest associations at the neighbourhood level.

Results from unadjusted models are not presented, but overall displayed similar associations to the adjusted models.

## Discussion

In this retrospective study of 26,152 confirmed cases of COVID-19 residing in 21,226 private households, we found that longer testing delays and male sex were associated with greater odds of household secondary transmission, while being a healthcare worker or linked to a known outbreak was associated with lower odds of household transmission. Additionally, neighbourhoods with larger average economic family size and a higher proportion of households with multiple persons per room were associated with greater odds of household transmission.

Previous studies of household transmission have considered secondary attack rates (SARs), defined as the proportion of household members of confirmed cases that acquire infection. The majority of these studies were conducted in Asia, and some in Europe and the United States.^4–6,8–10^ Madewell et al.^4^, Lei et al.^6^, and Curmei et al.^5^ conducted meta-analyses of previous studies and found pooled household SARs of 19% (95% CI: 15% - 23%), 27% (21% - 32%), and 30% (18% - 43%), respectively. Some of the included studies compared SARs in household settings verses non-household settings, and pooled estimates found that household SARs were five^4^ to ten^6^ times as high as non-household SARs, which highlights the role of household transmission in the spread of COVID-19.

We identified only two other studies that considered the impact of testing delays on household transmission; Xin et al.^17^ and Wang et al.^14^ examined the time from illness onset to laboratory confirmation. They reported hazard ratios for household transmission of 2·32 (95% CI 0·89 - 6·10) (<7-day delays versus ≥7-day delays) calculated from 106 households, and 2·35 (95% CI 0·63 - 8·77) (<3-day delays versus ≥3-day delays) calculated from 124 households, respectively. It has been estimated that infectivity peaks 3-5 days after symptom onset^18,19^, which underlines the importance of rapid testing and self-isolation as soon as symptoms appear. Our other finding of lower odds of household transmission among individuals with no symptoms is in line with estimates of lower SARs among asymptomatic or mildly symptomatic individuals^4,20^, although it may be that SARs are underestimated in these groups due to lower testing rates.^20^ Our “no symptom” classification may have included some individuals who missed having their symptoms reported in provincial disease systems, however we would expect these individuals to bias our estimate towards the null.

Considering other individual-level characteristics, two studies found similar positive associations with male sex and immunodeficiency^8^, and an inverse association with healthcare employment.^21^ In addition to healthcare employment, we also found lower odds of household transmission among individuals linked to a known outbreak. This may reflect testing practices, where outbreak-linked cases are identified and isolated faster than non-outbreak-linked cases. Healthcare workers may also be part of these outbreaks, leading to more rapid identification; additionally, they may have different practices within the household given their heightened awareness of risk of exposure, and may have increased access to or use of personal protective equipment as compared to non-healthcare workers.

Madewell et al.^4^ and Lopez Bernal et al.^10^ further reported inverse relationships between household size and SAR. These findings are contrasted to our result of higher odds of household transmission among neighbourhoods with larger average economic family size. Madewell et al. acknowledged that household crowding may play a more important role in transmission risk than household size; Lewis et al.^8^ found a relative risk of 2·1 (95% CI: 1·5 - 2·8) for transmission in households with >2 persons per bedroom compared to 1-2 persons per bedroom. Our findings of higher odds of household transmission among neighbourhoods with a higher proportion of multiple persons per room and multi-family households may support this hypothesis, and our association with economic family size may be capturing aspects of household crowding at the neighbourhood-level (e.g., neighbourhoods with larger average economic family size tended to be neighbourhoods with a higher proportion of multiple persons per room). One approach that has been implemented in some jurisdictions to mitigate this issue is voluntary self-isolation facilities for those who are unable to self-isolate in their home. Madewell et al. also reported a pooled proportion of households with any secondary transmission of 33% (95% CI 7% - 58%), while we found only 14% of our included households experienced secondary transmission. As we did not have information on total household size, it may be that we included some single-resident households that had zero probability of household transmission. This would decrease the number of cases associated with household transmission in comparison to studies that excluded single-resident households, and may have also diluted our model estimates.

Our study has some limitations that merit discussion. First, we did not have information on the total number of individuals residing in each household or the characteristics of uninfected household members, thus we were unable to calculate the proportion of household contacts infected to generate SARs. However, we were able to control for some neighbourhood-level characteristics of household composition including economic family size and proportion of households with multiple persons per room or multi-family households. Our finding of high transmission and acquisition of SARS-CoV-2 infection between individuals in the same age group therefore likely reflects the inherent age structures of households in Ontario. Second, we may have misclassified some index cases if a previously infected individual within the household was untested (e.g., asymptomatic or symptomatic but did not seek testing), and we may have misclassified some secondary cases if their infection was acquired outside the household. We may also have missed secondary cases within a household that were untested. Third, we only considered one index case per household, and considered all subsequent cases within a 14-day period to be secondary cases (i.e., did not account for tertiary transmission). Fourth, as this study encompasses a period before schools were re-opened in the fall, there were few index cases among children (N=190) and as such, we were not able to determine the extent to which children played a role in household transmission. Finally, because addresses in this dataset are entered manually as a free-text field, some algorithm misclassification is expected due to incorrectly entered addresses or different street and city naming conventions. This type of misclassification would be expected to decrease our pool of multiple-case households.

Our study also has several strengths. To our knowledge, this study contains the largest number of private households with at least one confirmed case of COVID-19. Most previous studies included a subset of confirmed COVID-19 cases, and used contact tracing to monitor household members for infection and/or symptoms.^4^ Thus, these studies were only able to include a smaller number of households (individual studies reporting on fewer than 6000 households) compared to the 21,226 households we were able to identify through address matching of all confirmed cases of COVID-19 in Ontario. We did not find any other studies that used address matching to comprehensively identify all households with SARS-CoV-2 infections in a region, with the exception of one study from Israel that used a municipal database of residents to identify household members of cases.^22^ Additionally, we considered a large set of individual- and neighbourhood-level characteristics of index cases. We were able to compare these characteristics between households where secondary transmission did and did not occur, which yielded important insights into factors that may help reduce secondary transmission in households.

## Conclusion

Household transmission plays a key role in local spread of SARS-CoV-2 infection. Our work suggests that it is important for individuals to get tested for COVID-19 as soon as symptoms appear. Ideally, individuals should be tested on the day of symptom onset, as even a 1-day delay was associated with increased odds of secondary transmission. Additionally, if cases are living with other individuals, it may also be important to try to isolate in a room alone or outside the home, if possible. These strategies may be considered by public health officials to reduce household transmission and help mitigate the ongoing spread of COVID-19. Future research should focus on the role of children and youth in household transmission, particularly as lockdown restrictions are lifted and individuals return to regular activities such as work, school, and daycare.

## Data Availability

Data sharing requests should be directed to Public Health Ontario.

## Acknowledgements

The authors would like to thank Garima Aryal for her contributions to the address matching work.

## Notes

**Declaration of interests** The authors have no conflicts of interest to declare.

### Competing Interest Statement

The authors have declared no competing interest.

### Author Declarations

We obtained ethics approval from Public Health Ontario's Research Ethics Board.

