## Supplementary Material for "Characteristics associated with household transmission of SARS-CoV-2 in Ontario, Canada"

*Affiliations:*
^a^Health Protection, Public Health Ontario, 661 University Ave., Floor 17, Toronto, ON, M5G 1M1, Canada
^b^Sunnybrook Research Institute, Sunnybrook Health Sciences Centre, 2075 Bayview Ave., Toronto, ON, M4N 3M5, Canada
^c^ICES, 2075 Bayview Ave., Room G1 06, Toronto, ON, M4N 3M5, Canada
^d^Division of Infectious Diseases, Sunnybrook Health Sciences Centre, 2075 Bayview Ave., Room B1 03, Toronto, ON, M4N 3M5, Canada

^e^Department of Medicine, University of Toronto, 6 Queen’s Park Cres. W, Floor 3, Toronto, ON, M5S 3H2, Canada

^f^Institute of Health Policy, Management and Evaluation, University of Toronto, 155 College St., Suite 425, Toronto, ON, M5T 3M6, Canada

^g^Dalla Lana School of Public Health, University of Toronto, 155 College St., Room 500, Toronto, ON, M5T 3M7, Canada
^h^Environmental and Occupational Health, Public Health Ontario, 480 University Ave., Suite 300, Toronto, ON, M5G 1V2, Canada
^i^Knowledge Services, Public Health Ontario, 480 University Ave., Suite 300, Toronto, ON, M5G 1V2, Canada

*Corresponding Author:*
Sarah Buchan, PhD
Public Health Ontario
661 University Avenue, Floor 17
Toronto, Ontario, M5G 1M1


### **Supplementary Definitions**

**Housing suitability:** Private dwellings that have enough bedrooms for the size and composition of the household. Based on the age, sex, and relationships among household members.^1^

**Multiple persons per room:** An indicator of the level of crowding in a private dwelling. Calculated by dividing the number of persons in the household by the number of rooms in the dwelling.^1^

**Census family:** A married couple and the children, if any, of either and/or both spouses; a couple living common law and the children, if any, of either and/or both partners; or a lone parent of any marital status with at least one child living in the same dwelling and that child or those children. All members of a particular census family live in the same dwelling.^1^

**Multi-family households:** Households in which two or more census families occupy the same private dwelling.^1^

**Economic family:** A group of two or more persons who live in the same dwelling and are related to each other by blood, marriage, common-law union, adoption or a foster relationship. By definition, all persons who are members of a census family are also members of an economic family.^1^

**Community type:** Categorized using Health Quality Ontario’s geographic stratifier approach, which groups dissemination areas into a four category urban/rural continuum based on community size, population density, and level of integration with a census metropolitan area or census agglomeration.^2^

### **Supplementary Tables**

**Supplementary Table S1.** Health regions and public health units across Ontario

| **Health Region** | **Public Health Units** |
| --- | --- |
| North | Northwestern Health Unit, Thunder Bay District Health Unit, Algoma Public Health, North Bay Parry Sound District Health Unit, Porcupine Health Unit, Public Health Sudbury & Districts, and Timiskaming Health Unit |
| East | Ottawa Public Health, Eastern Ontario Health Unit, Hastings Prince Edward Public Health, Kingston, Frontenac and Lennox & Addington Public Health, Leeds, Grenville & Lanark District Health Unit, and Renfrew County and District Health Unit |
| Central East | Durham Region Health Department, Haliburton, Kawartha, Pine Ridge District Health Unit, Peel Public Health, Peterborough Public Health, Simcoe Muskoka District Health Unit, and York Region Public Health |
| Central West | Brant County Health Unit, City of Hamilton Public Health Services, Haldimand-Norfolk Health Unit, Halton Region Public Health, Niagara Region Public Health, Region of Waterloo Public Health and Emergency Services, and Wellington-Dufferin-Guelph Public Health |
| Toronto | Toronto Public Health |
| South West | Chatham-Kent Public Health, Grey Bruce Health Unit, Huron Perth Public Health, Lambton Public Health, Middlesex-London Health Unit, Southwestern Public Health, and Windsor-Essex County Health Unit |

**Supplementary Table S2.** Characteristics of index cases with household transmission compared to secondary cases

|  | **Index cases with**  **household transmission**  **(N=3,067)** | **Secondary cases**  **(N=4,656)** |
| --- | --- | --- |
| Sex (N, %) |  |  |
| Female | 1,464 (47·7) | 2,378 (51·1) |
| Male | 1,595 (52·0) | 2,270 (48·8) |
| Age, years (median, IQR) | 46 [31, 57] | 38 [23, 55] |
| Age group (N, %) |  |  |
| <10 years | 26 (0·8) | 270 (5·8) |
| 10-19 years | 127 (4·1) | 466 (10·0) |
| 20-29 years | 523 (17·1) | 1,044 (22·4) |
| 30-39 years | 481 (15·7) | 667 (14·3) |
| 40-49 years | 571 (18·6) | 615 (13·2) |
| 50-59 years | 726 (23·7) | 772 (16·6) |
| 60-69 years | 404 (13·2) | 499 (10·7) |
| 70-79 years | 138 (4·5) | 221 (4·7) |
| ≥80 years | 70 (2·3) | 102 (2·2) |
| High risk (≥60 years, immunocompromised, cardiovascular, COPD) (N, %) | 844 (27·5) | 1,036 (22·3) |
| Outbreak-associated* (N, %) | 540 (17·6) | 360 (7·7) |
| Healthcare worker (N, %) | 517 (16·9) | 397 (8·5) |
| Month reported (N, %) |  |  |
| January | 1 (0·0) | 1 (0·0) |
| February | 3 (0·1) | 3 (0·1) |
| March | 312 (10·2) | 273 (5·9) |
| April | 945 (30·8) | 1,268 (27·2) |
| May | 989 (32·2) | 1,596 (34·3) |
| June | 528 (17·2) | 997 (21·4) |
| July | 289 (9·4) | 518 (11·1) |
| Region (N, %) |  |  |
| Toronto | 1,025 (33·4) | 1,548 (33·2) |
| Central East | 1,236 (40·3) | 1,948 (41·8) |
| Central West | 343 (11·2) | 508 (10·9) |
| Eastern | 231 (7·5) | 314 (6·7) |
| Northern | 32 (1·0) | 40 (0·9) |
| South West | 200 (6·5) | 298 (6·4) |
| Testing delay†, days (median, IQR) | 4 [2, 8] | 2 [0, 4] |
| Testing delay distribution† (N, %) |  |  |
| No symptoms‡ | 131 (4·3) | 751 (16·3) |
| <0 days§ | 60 (2·0) | 309 (6·7) |
| 0 days | 164 (5·4) | 539 (11·7) |
| 1 day | 349 (11·5) | 580 (12·6) |
| 2 days | 341 (11·2) | 521 (11·3) |
| 3 days | 327 (10·8) | 431 (9·4) |
| 4 days | 276 (9·1) | 320 (7·0) |
| ≥5 days | 1,390 (45·8) | 1,146 (24·9) |
| Reporting delay, days (median, IQR) | 2 [1, 3] | 2 [1, 3] |
| Reporting delay distribution (N, %) |  |  |
| <0 days | 43 (1·4) | 98 (2·1) |
| 0 days | 200 (6·5) | 301 (6·5) |
| 1 day | 1,038 (34·0) | 1,604 (34·5) |
| 2 days | 926 (30·3) | 1,419 (30·6) |
| 3 days | 390 (12·8) | 612 (13·2) |
| 4 days | 188 (6·2) | 237 (5·1) |
| ≥5 days | 271 (8·9) | 373 (8·0) |
| Data entry delay, days (median, IQR) | 0 [0, 1] | 0 [0, 1] |
| Data entry delay distribution (N, %) |  |  |
| <0 days | 173 (5·6) | 580 (12·5) |
| 0 days | 1,852 (60·4) | 2,593 (55·7) |
| 1 day | 696 (22·7) | 1,003 (21·5) |
| 2 days | 132 (4·3) | 196 (4·2) |
| 3 days | 80 (2·6) | 89 (1·9) |
| 4 days | 32 (1·0) | 66 (1·4) |
| ≥5 days | 102 (3·3) | 129 (2·8) |

*Cases associated with a public health declared outbreak outside the home.
†65 cases excluded that were missing symptom onset date but had COVID-19 symptoms flagged in provincial reportable disease systems.
‡Cases with no symptoms were defined as cases that were missing symptom onset date (thus specimen collection date was used) and did not have any COVID-19 symptoms flagged in provincial reportable disease systems.
§Cases with a testing delay of <0 days were those who were tested prior to the onset of their symptoms.

**Supplementary Table S3.** Outcome severity of index cases, secondary cases, and cases that were not involved in any household transmission

|  | **No serious or severe outcome (N=22,884)** | **Serious or severe outcome***  **(N=2,998)** |
| --- | --- | --- |
| Cases with no transmission (N, row %) | 15,907 (87·6) | 2,252 (12·4) |
| Index cases (N, row %) | 2,689 (87·7) | 378 (12·3) |
| Secondary cases (N, row %) | 4,288 (92·1) | 368 (7·9) |
| **Secondary cases by age group of index case†** | | |
| Age group (N, row %) |  |  |
| <10 years | 34 (91·9) | 3 (8·1) |
| 10-19 years | 232 (95·5) | 11 (4·5) |
| 20-29 years | 810 (95·1) | 42 (4·9) |
| 30-39 years | 725 (94·6) | 41 (5·4) |
| 40-49 years | 779 (92·0) | 68 (8·0) |
| 50-59 years | 979 (92·2) | 83 (7·8) |
| 60-69 years | 509 (89·5) | 60 (10·5) |
| 70-79 years | 162 (85·3) | 28 (14·7) |
| ≥80 years | 57 (64·0) | 32 (36·0) |

*Serious outcomes included any hospitalization and severe outcomes included ICU admission or death.
†One secondary case with no serious or severe outcome had an index case with missing age, therefore this column only totals to 4,287.

**Supplementary Table S4.** Adjusted odds ratios and 95% confidence intervals for the associations between index case delay metrics and odds of household transmission

|  | **OR (95% CI)*** | | |
| --- | --- | --- | --- |
|  | **Any household transmission** | **Household transmission**  **to older adults**  **(aged** ≥**60 years)** | **Household transmission to severe cases**  **(ICU or death)** |
| Testing delay† |  |  |  |
| No symptoms‡ | 0·48 (0·38 - 0·61) | 0·38 (0·23 - 0·62) | 0·49 (0·19 - 1·28) |
| <0 days§ | 0·69 (0·51 - 0·94) | 0·83 (0·47 - 1·45) | 0·17 (0·02 - 1·31) |
| 0 days | Ref | Ref | Ref |
| 1 day | 2·02 (1·66 - 2·48) | 2·02 (1·38 - 2·96) | 0·44 (0·15 - 1·28) |
| 2 days | 1·96 (1·60 - 2·40) | 1·48 (0·99 - 2·22) | 1·25 (0·56 - 2·78) |
| 3 days | 2·36 (1·93 - 2·90) | 2·49 (1·70 - 3·65) | 1·90 (0·88 - 4·09) |
| 4 days | 2·64 (2·14 - 3·26) | 2·54 (1·70 - 3·80) | 1·59 (0·68 - 3·71) |
| ≥5 days | 3·02 (2·53 - 3·60) | 2·88 (2·07 - 4·00) | 2·28 (1·20 - 4·32) |
| Reporting delay |  |  |  |
| <0 days | 0·82 (0·57 - 1·17) | 0·70 (0·36 - 1·33) | 1·54 (0·28 - 8·49) |
| 0 days | Ref | Ref | Ref |
| 1 day | 0·97 (0·82 - 1·15) | 0·90 (0·66 - 1·22) | 3·13 (1·13 - 8·67) |
| 2 days | 0·98 (0·83 - 1·17) | 0·95 (0·69 - 1·30) | 2·90 (1·03 - 8·12) |
| 3 days | 0·95 (0·78 - 1·15) | 0·98 (0·69 - 1·38) | 2·66 (0·89 - 7·96) |
| 4 days | 1·09 (0·87 - 1·36) | 1·08 (0·72 - 1·61) | 2·92 (0·89 - 9·55) |
| ≥5 days | 0·73 (0·59 - 0·89) | 0·77 (0·54 - 1·11) | 2·04 (0·67 - 6·25) |
| Data entry delay |  |  |  |
| <0 days | 0·94 (0·78 - 1·12) | 0·90 (0·63 - 1·27) | 0·71 (0·30 - 1·69) |
| 0 days | Ref | Ref | Ref |
| 1 day | 1·04 (0·94 - 1·15) | 0·87 (0·72 - 1·05) | 0·87 (0·57 - 1·32) |
| 2 days | 0·92 (0·76 - 1·12) | 0·99 (0·70 - 1·41) | 1·36 (0·72 - 2·56) |
| 3 days | 1·04 (0·81 - 1·34) | 0·66 (0·38 - 1·15) | 0·37 (0·09 - 1·54) |
| 4 days | 0·62 (0·42 - 0·91) | 0·62 (0·30 - 1·27) | 0·58 (0·14 - 2·42) |
| ≥5 days | 0·75 (0·60 - 0·93) | 0·73 (0·48 - 1·12) | 1·04 (0·50 - 2·16) |

*Estimates were adjusted for age group, sex, month reported, health region, and economic family size.
†269 cases were excluded from the testing delay models that had COVID-19 symptoms flagged in provincial reportable disease systems but were missing symptom onset date.
‡Cases with no symptoms were defined as cases that were missing symptom onset date (thus specimen collection date was used) and did not have any COVID-19 symptoms flagged in provincial reportable disease systems.
§Cases with a testing delay of <0 days were those who were tested prior to the onset of their symptoms.

**Supplementary Table S5.** Sensitivity analyses for the associations between index case characteristics and odds of any household transmission

|  | **OR (95% CI)*** | | |
| --- | --- | --- | --- |
|  | **Secondary cases occurring 2-14 days after index** | **Secondary cases occurring 1-28 days after index** | **Households with index dates on or after May 29** |
| **Individual-level characteristics** | | | |
| Testing delay† |  |  |  |
| No symptoms‡ | 0·51 (0·39 - 0·67) | 0·50 (0·40 - 0·63) | 0·47 (0·32 - 0·69) |
| <0 days§ | 0·77 (0·55 - 1·07) | 0·69 (0·51 - 0·93) | 0·86 (0·53 - 1·40) |
| 0 days | Ref | Ref | Ref |
| 1 day | 2·17 (1·74 - 2·71) | 1·99 (1·64 - 2·42) | 2·36 (1·66 - 3·36) |
| 2 days | 2·21 (1·77 - 2·75) | 1·99 (1·64 - 2·42) | 2·11 (1·47 - 3·04) |
| 3 days | 2·61 (2·09 - 3·26) | 2·25 (1·85 - 2·75) | 2·31 (1·58 - 3·37) |
| 4 days | 2·96 (2·35 - 3·72) | 2·50 (2·03 - 3·07) | 2·76 (1·85 - 4·12) |
| ≥5 days | 3·48 (2·87 - 4·22) | 3·13 (2·64 - 3·71) | 3·01 (2·17 - 4·19) |
| Reporting delay |  |  |  |
| <0 days | 0·74 (0·50 - 1·10) | 0·94 (0·67 - 1·31) | 1·26 (0·32 - 4·88) |
| 0 days | Ref | Ref | Ref |
| 1 day | 1·00 (0·84 - 1·20) | 0·95 (0·80 - 1·12) | 1·17 (0·72 - 1·91) |
| 2 days | 0·99 (0·83 - 1·19) | 0·97 (0·82 - 1·14) | 1·14 (0·70 - 1·85) |
| 3 days | 0·97 (0·79 - 1·19) | 0·94 (0·78 - 1·13) | 1·17 (0·71 - 1·95) |
| 4 days | 1·12 (0·89 - 1·42) | 1·12 (0·90 - 1·38) | 0·93 (0·51 - 1·71) |
| ≥5 days | 0·76 (0·62 - 0·95) | 0·73 (0·60 - 0·89) | 0·46 (0·21 - 1·02) |
| Data entry delay |  |  |  |
| <0 days | 0·94 (0·78 - 1·14) | 0·92 (0·77 - 1·09) | 0·90 (0·65 - 1·25) |
| 0 days | Ref | Ref | Ref |
| 1 day | 1·03 (0·93 - 1·14) | 1·04 (0·95 - 1·15) | 1·25 (1·01 - 1·54) |
| 2 days | 0·97 (0·79 - 1·19) | 0·94 (0·78 - 1·14) | 1·75 (0·75 - 4·09) |
| 3 days | 1·04 (0·80 - 1·36) | 1·06 (0·83 - 1·35) | Insufficient data |
| 4 days | 0·61 (0·41 - 0·92) | 0·69 (0·48 - 0·97) | Insufficient data |
| ≥5 days | 0·77 (0·61 - 0·97) | 0·75 (0·61 - 0·93) | 0·50 (0·11 - 2·23) |
| Sex |  |  |  |
| Female | Ref | Ref | Ref |
| Male | 1·32 (1·21 - 1·43) | 1·29 (1·19 - 1·39) | 1·15 (0·97 - 1·35) |
| Age group |  |  |  |
| <10 years | 0·82 (0·52 - 1·30) | 0·81 (0·53 - 1·24) | 1·20 (0·68 - 2·11) |
| 10-19 years | 1·18 (0·94 - 1·47) | 1·19 (0·96 - 1·47) | 1·17 (0·81 - 1·67) |
| 20-29 years | 0·78 (0·68 - 0·89) | 0·79 (0·70 - 0·89) | 0·91 (0·71 - 1·16) |
| 30-39 years | 0·80 (0·70 - 0·92) | 0·80 (0·71 - 0·91) | 0·87 (0·66 - 1·14) |
| 40-49 years | 0·92 (0·81 - 1·05) | 0·90 (0·80 - 1·01) | 1·00 (0·75 - 1·32) |
| 50-59 years | Ref | Ref | Ref |
| 60-69 years | 0·93 (0·81 - 1·07) | 0·94 (0·82 - 1·07) | 0·95 (0·70 - 1·30) |
| 70-79 years | 0·79 (0·64 - 0·98) | 0·78 (0·64 - 0·94) | 0·98 (0·63 - 1·52) |
| ≥80 years | 0·56 (0·42 - 0·74) | 0·61 (0·48 - 0·79) | 0·72 (0·38 - 1·36) |
| High risk (≥60 years, immunocompromised, cardiovascular, COPD) |  |  |  |
| No | Ref | Ref | Ref |
| Yes | 1·09 (0·93 - 1·29) | 1·13 (0·97 - 1·32) | 0·96 (0·65 - 1·41) |
| Outbreak-associated^\|\|^ |  |  |  |
| No | Ref | Ref | Ref |
| Yes | 0·64 (0·57 - 0·71) | 0·63 (0·57 - 0·69) | 0·76 (0·60 - 0·97) |
| Healthcare worker |  |  |  |
| No | Ref | Ref | Ref |
| Yes | 0·57 (0·51 - 0·64) | 0·56 (0·51 - 0·63) | 0·55 (0·40 - 0·74) |
| Month reported |  |  |  |
| January | Insufficient data | 7·10 (0·41 - 122·92) | NA |
| February | 2·94 (0·77 - 11·29) | 2·29 (0·60 - 8·77) | NA |
| March | 1·06 (0·91 - 1·23) | 1·04 (0·91 - 1·20) | NA |
| April | Ref | Ref | NA |
| May | 1·22 (1·10 - 1·36) | 1·23 (1·12 - 1·35) | Ref |
| June | 1·15 (1·02 - 1·31) | 1·11 (0·99 - 1·25) | 1·30 (0·72 - 2·37) |
| July | 1·03 (0·88 - 1·20) | 0·98 (0·85 - 1·13) | 1·35 (0·74 - 2·47) |
| Region |  |  |  |
| Toronto | Ref | Ref | Ref |
| Central East | 1·02 (0·92 - 1·13) | 1·02 (0·92 - 1·12) | 0·99 (0·81 - 1·22) |
| Central West | 0·92 (0·79 - 1·07) | 0·89 (0·78 - 1·02) | 0·71 (0·52 - 0·97) |
| Eastern | 1·09 (0·93 - 1·29) | 1·06 (0·91 - 1·23) | 0·88 (0·63 - 1·22) |
| Northern | 1·15 (0·75 - 1·77) | 1·31 (0·90 - 1·91) | 0·54 (0·16 - 1·79) |
| South West | 0·84 (0·70 - 1·00) | 0·83 (0·71 - 0·98) | 0·65 (0·48 - 0·88) |
| **Neighbourhood-level characteristics**¶ | | | |
| % Age group |  |  |  |
| 0-14 years | 0·86 (0·75 - 1·00) | 0·88 (0·77 - 1·00) | 0·99 (0·75 - 1·30) |
| 15-64 years | 0·96 (0·86 - 1·06) | 0·94 (0·85 - 1·03) | 0·93 (0·76 - 1·14) |
| ≥65 years | 1·11 (1·01 - 1·22) | 1·11 (1·02 - 1·21) | 1·07 (0·89 - 1·28) |
| % Male | 0·84 (0·63 - 1·11) | 0·91 (0·70 - 1·17) | 1·09 (0·64 - 1·85) |
| % Recent immigrants | 1·34 (1·20 - 1·51) | 1·36 (1·23 - 1·52) | 1·46 (1·17 - 1·82) |
| % Non-White, non-Indigenous | 1·05 (1·02 - 1·08) | 1·06 (1·03 - 1·08) | 1·10 (1·04 - 1·15) |
| Non-White, non-Indigenous groups |  |  |  |
| % Black | 1·04 (0·98 - 1·10) | 1·07 (1·01 - 1·12) | 1·11 (1·00 - 1·24) |
| % East/Southeast Asian | 1·03 (0·99 - 1·07) | 1·02 (0·98 - 1·05) | 1·04 (0·97 - 1·12) |
| % Latin American | 1·41 (1·18 - 1·68) | 1·46 (1·24 - 1·72) | 1·32 (0·95 - 1·84) |
| % Middle Eastern | 1·07 (0·97 - 1·19) | 1·05 (0·95 - 1·15) | 1·21 (1·00 - 1·47) |
| % South Asian | 1·04 (1·00 - 1·07) | 1·04 (1·01 - 1·08) | 1·06 (0·99 - 1·14) |
| % Below low income cut-off | 1·07 (1·00 - 1·14) | 1·07 (1·01 - 1·13) | 1·20 (1·07 - 1·34) |
| % Labour force participation | 0·90 (0·83 - 0·96) | 0·89 (0·83 - 0·95) | 0·84 (0·73 - 0·97) |
| % Less than high school education | 1·09 (1·03 - 1·17) | 1·10 (1·03 - 1·16) | 1·12 (0·99 - 1·26) |
| % Unsuitable housing# | 1·18 (1·11 - 1·27) | 1·20 (1·13 - 1·28) | 1·24 (1·09 - 1·41) |
| % Households with multiple persons per room# | 1·23 (1·11 - 1·37) | 1·26 (1·14 - 1·39) | 1·34 (1·10 - 1·62) |
| % Multi-family households# | 1·15 (0·96 - 1·38) | 1·15 (0·97 - 1·36) | 0·99 (0·70 - 1·41) |
| Economic family size# | 1·92 (1·72 - 2·15) | 1·96 (1·77 - 2·17) | 1·90 (1·54 - 2·35) |
| % Households living in apartments with ≥5 floors | 1·02 (1·00 - 1·05) | 1·02 (1·00 - 1·04) | 1·05 (1·01 - 1·09) |
| % Households living in apartments with <5 floors | 1·01 (0·96 - 1·05) | 1·01 (0·97 - 1·05) | 0·99 (0·90 - 1·08) |
| Community type# |  |  |  |
| Large urban | Ref | Ref | Ref |
| Medium/small | 0·91 (0·76 - 1·09) | 0·95 (0·81 - 1·12) | 0·57 (0·39 - 0·85) |
| Rural | 0·94 (0·76 - 1·17) | 0·99 (0·81 - 1·21) | 0·63 (0·40 - 1·00) |
| Remote | 0·77 (0·55 - 1·09) | 0·72 (0·52 - 0·99) | 0·54 (0·25 - 1·15) |

*Estimates were adjusted for age group, sex, month reported, health region, and economic family size.
†Cases were excluded from the testing delay models that had COVID-19 symptoms flagged in provincial reportable disease systems but were missing symptom onset date.
‡Cases with no symptoms were defined as cases that were missing symptom onset date (thus specimen collection date was used) and did not have any COVID-19 symptoms flagged in provincial reportable disease systems.
§Cases with a testing delay of <0 days were those who were tested prior to the onset of their symptoms.
^||^Cases associated with a public health declared outbreak outside the home.
¶Odds ratios for neighbourhood-level characteristics are reported per 10% increase, except for economic family size and community type.
#Defined in Supplementary Definitions.
NA=not applicable.

### **Supplementary Figures**

**Supplementary Figure S1.** Distribution of serial intervals (number of days between symptom onset dates for index cases and secondary cases)

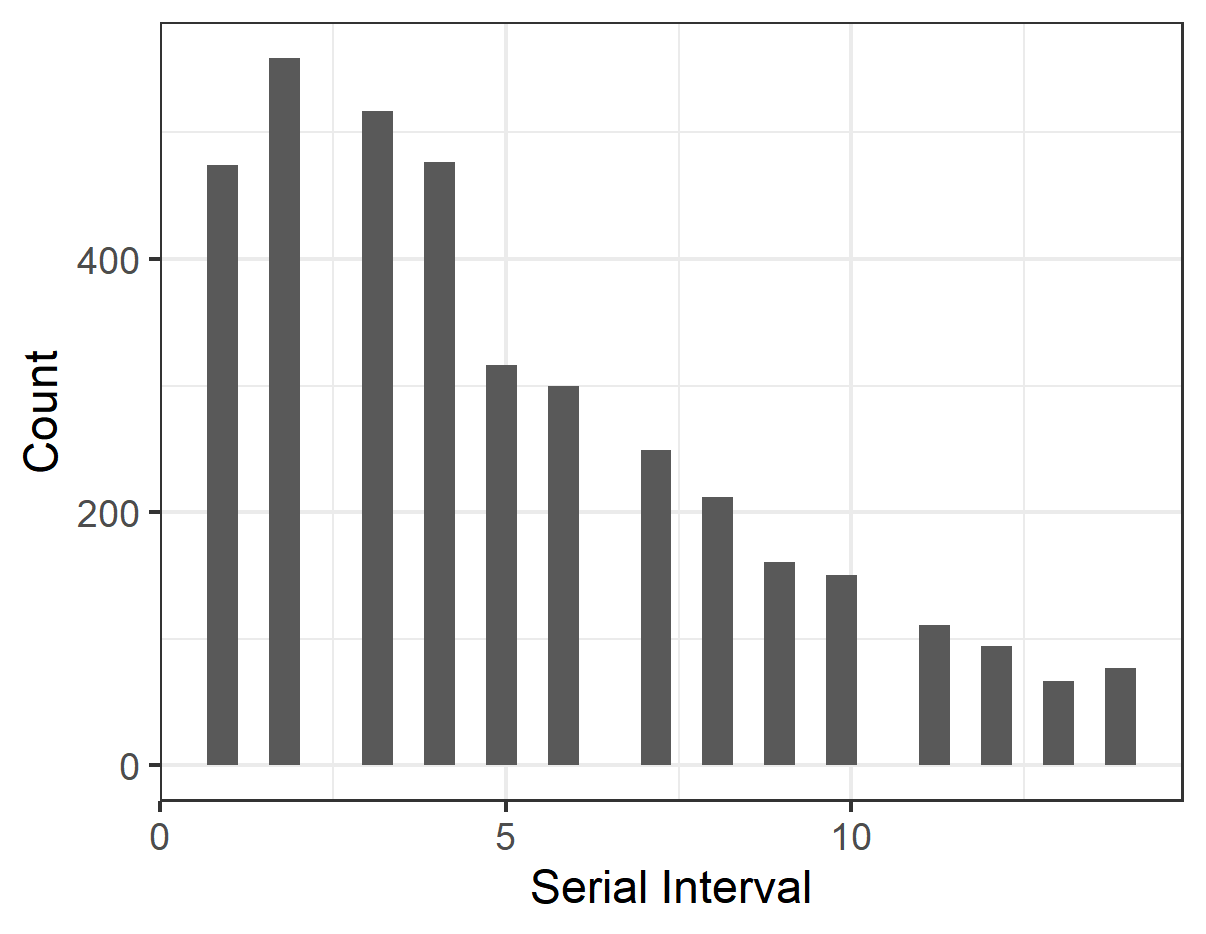
